# Pharmacokinetics of anti-SARS-CoV-2 monoclonal antibody SA55 injection in healthy participants

**DOI:** 10.1101/2025.02.22.25322730

**Authors:** Yibo Zhou, Xing Meng, Jianhua Li, Jin Wang, Gang Zeng, Yunlong Cao, Chaoying Hu, Ronghua Jin

## Abstract

To explore novel treatment/prevention of COVID-19, a novel broad-spectrum neutralizing antibody injection, anti-SARS-CoV-2 antibody SA55 Injection (SA55 injection) was developed. Pharmacokinetics (PK) characteristics of SA55 injection were evaluated in a randomized, controlled, double-blind phase □ trial based on healthy participants aged 18-65 years. PK parameters (AUC_0-∞_, AUC_0-t_, and C_max_) were assessed using one-way ANOVA and the Power model. Results demonstrated that SA55 injection with a T_max_ of 12.6 days and a half-life of 103 days. In conclusion, the SA55 injection demonstrates potential for use in the prevention of COVID-19 infection.

## Introduction

Starting from the end of 2019, a novel acute respiratory infection disease rapidly spread worldwide, affecting multiple countries. On January 31, 2020, the World Health Organization (WHO) declared the pneumonia outbreak caused by the novel coronavirus a Public Health Emergency of International Concern (PHEIC)^[1]^. On March 11, 2020, the WHO declared that the outbreak had entered a phase of international pandemic^[2]^. On February 12, 2020, the International Committee on Taxonomy of Viruses (ICTV) announced the official classification name of the novel coronavirus as Severe Acute Respiratory Syndrome Coronavirus 2 (SARS-CoV-2), while the WHO simultaneously announced the official name of the disease caused by this virus as COVID-19^[3]^.

During the COVID-19 pandemic, various vaccines and therapies for COVID-19 were widely implemented, making significant contributions to the prevention and control of the disease^[4, 5]^ The continuous evolution and emergence of SARS-CoV-2 variants pose significant challenges to controlling the global pandemic and raise concerns about the efficacy of monoclonal antibody therapies and vaccines^[6, 7]^. Neutralizing antibody (NAb) drugs against SARS-CoV-2 have shown good efficacy in preventing or treating COVID-19 by directly binding to the virus, thereby inhibiting further infection. According to data published by COVID-NMA, as of October 16, 2022, there are more than 370 investigational drugs for the novel coronavirus globally^[8]^. The U.S. Food and Drug Administration (FDA) has authorized the emergency use of several SARS-CoV-2 antibody therapies during the pandemic, including monotherapies such as tocilizumab (Roche), 1% Propofol-Lipuro injection (B. Braun), bamlanivimab (Lilly)^[9]^, and sotrovimab (GSK)^[10]^; as well as combination therapies like Evusheld (Tixagevimab/Cilgavimab) (AstraZeneca)^[11]^, REGEN-COV (Regeneron)^[12, 13]^, and bamlanivimab/etesevimab (Lilly)^[14, 15]^. According to incomplete statistics, there are currently over 80 candidate drugs for COVID-19 in China approved to initiate clinical trials, with 12 candidate drugs having entered the clinical stage. Among domestic drugs, the combination therapy of bimagrumab and romlusevimab by Brii Biosciences was conditionally approved for marketing by the National Medical Products Administration (NMPA) in December 2021.6^[16]^.

With the continuous mutation of the virus, many of the currently available neutralizing antibodies do not meet the needs for COVID-19 prevention and treatment^[17]^. Due to the rapid mutation of the spike protein of the novel coronavirus, and since neutralizing antibodies are designed to target the structure of the virus’s spike protein on cells, the use of neutralizing antibodies is limited by the sensitivity of circulating strains to these antibodies^[18]^. This has led to the emergence of many variants, posing new challenges for COVID-19^[7, 19-23]^. With the global prevalence of the Omicron variant, the in vitro activity of various neutralizing antibodies against the Omicron strain has rapidly diminished^[7]^. New variants have the ability to evade antibodies obtained through vaccination or previous SARS-CoV-2 infections, as well as neutralizing antibody (NAb) drugs targeting SARS-CoV-2^[24]^. Currently, the monoclonal antibody treatments for COVID-19 approved for Emergency Use Authorization (EUA) target the receptor-binding domain (RBD) of the SARS-CoV-2 spike protein. Although monoclonal antibody combinations (antibody cocktail therapies^[25-27]^have been developed to prevent potential neutralization escape by targeting multiple viral epitopes, many neutralizing antibody treatments are still evaded by variants of the novel coronavirus^[20, 21, 23, 28]^. Currently, the FDA has revoked all Emergency Use Authorizations for the previously issued COVID-19 neutralizing antibodies. Therefore, developing broad-spectrum neutralizing antibodies has become one of the crucial strategies for the global response to the rapid mutation of the novel coronavirus.

The ideal drug for treatment should have characteristics of rapid onset and strong potency, as well as broad-spectrum efficacy to address various variants of the novel coronavirus. To achieve these goals, a novel broad-spectrum neutralizing antibody injection, SA55, has been developed. The SA55 injection can bind to the novel coronavirus, thereby preventing the virus from entering host cells and blocking infection. The SA55 antibody was selected using an innovative high-throughput sequencing technology from a library of approximately 13,000 antibodies, which efficiently binds to conserved sites shared by various coronaviruses that are less prone to mutation^[24, 29]^. It effectively neutralized multiple Omicron variants, including BA.1, BA.2, BA.4/5, BF.7, XBB, BQ.1, and BQ.1.1, and is currently the only reported antibody drug globally that has not been evaded by the novel coronavirus^[30-35]^. Compared to existing therapies for COVID-19, this product possesses unique advantages and is expected to become an ideal choice for treating the novel coronavirus.

In this phase □ clinical trial, safety, tolerability, PK characteristics, immunogenicity, and serum neutralizing activity after intramuscular injection of SA55 injection were evaluated in healthy populations aged 18-65 years (ClinicalTrials.gov ID NCT06050460). In this article, PK characteristics of SA55 injection with 150mg dosage were reported.

## Objects and Methods

### Study Drugs

This study employs a placebo-controlled design, with both the investigational drug and placebo developed and produced by Sinovac Life Sciences Co., Ltd. The novel coronavirus broad-spectrum neutralizing antibody SA55 injection (Batch number: KC202301001A, Production date: January 3, 2023. Expiry date: January 2, 2025) contains the main component, the novel coronavirus broad-spectrum neutralizing antibody SA55. The placebo for the novel coronavirus broad-spectrum neutralizing antibody SA55 injection (Batch number: 20221228, Production date: December 29, 2022. Expiry date: December 28, 2022) does not contain neutralizing antibodies.

### Study Participants

The protocol and informed consent form were reviewed and approved by the ethics committee of the Beijing Ditan Hospital of Capital Medical University (NO. DTEC-YW2023-014-01) and the National Medical Products Administration of China before the start of the study. Each healthy participant gave written informed consent to participate in this study after being told of the objectives, procedures, and possible risks of the research. The study was conducted in accordance with the Declaration of Helsinki, Good Clinical Practice, and the relevant regulatory standards. The researchers screened the participants, including physical examinations, 12-lead electrocardiography, laboratory tests, with medical histories being either normal or only mildly abnormal without clinical significance.

The inclusion criteria were as follows: Participants recruited on the day of enrollment were males or females aged 18 to 65 years; male participants weighed ≥50.0 kg; female participants weighed ≥45.0 kg, and had a body mass index (BMI) between 18.0 and 28.0 kg/m^2^ (inclusive of boundary values). Exclusion criteria include: individuals with known allergies to the investigational drug or any components of the formulation, or to other similar drugs; potential interference at the target injection site (deltoid muscle of the upper arm) that may affect drug administration or local reaction observation; individuals who have received any marketed or investigational SARS-CoV-2 neutralizing antibody injection prior to screening; individuals with a known history of SARS-CoV-2 infection or vaccination against COVID-19 within the past three months.

### Study Design

This study employs a randomized, controlled, double-blind design, recruiting healthy participants aged 18 to 65 years. Ten participants were planned to be recruited in 150mg group (lowest dose group). After providing informed consent, participants were screened against the inclusion/exclusion criteria and sequentially enrolled from the lowest to the highest dose group. Participants in the lowest dose group received 150mg of the investigational drug or placebo.

In 150mg group, participants were randomly assigned in a 4:1 ratio to receive either the SA55 injection or an equivalent volume of placebo (with 8 participants receiving SA55 injection and 2 receiving placebo), with a “sentinel dosing” setup. Specifically, on the day of dosing, 2 participants were enrolled first and randomly assigned in a 1:1 ratio to receive either the SA55 injection or placebo. After monitoring for at least 24 hours and confirming the safety of the drug, the remaining 8 participants (with 7 receiving the SA55 injection and 1 receiving placebo) were then be enrolled and randomly assigned. All participants had blood samples collected at various time points (1, 2, 3, 5, 7, 11, 14, 21, 28, 56, 84, 112, 140, 168, 182 days after administratoin) during the study to measure drug concentrations. The test flowchart of 150mg group is shown in Figure 1.

**Figure 1.**
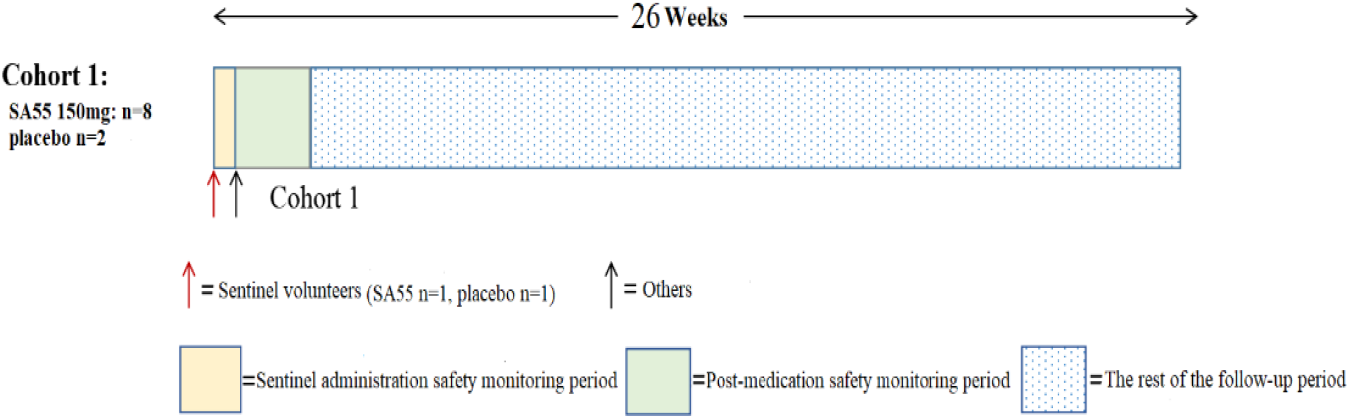
Study flowchart of 150mg group.

### Outcomes

The serum PK parameters of SA55 post-administration include T_max_, C_max_, elimination half-life (t_1/2_), clearance rate (CL), apparent volume of distribution (Vd), area under the drug concentration-time curve (AUC_0-t_, AUC_0-∞_), etc.

### Statistical Analysis

Continuous data were expressed as the means ± standard deviation (SD), and a one-way analysis of variance (ANOVA) was used to evaluate the dose proportionality of the PK parameters AUC_0-∞_, AUC_0-t_, and C_max_. All statistical analyses were performed using statistical software SAS 9.4 (SAS Institute, Cary, North Carolina) or later versions.

## Results

### Pharmacokinetic Properties

According to study protocol, a total of 10 participants in 150mg dose group completed the trial and were included in the PK analysis. The blood concentrations of SA55 injection were analyzed at various time points after administration based on 8 participants received SA55 injection. The average drug concentration-time plots after administration were shown in Figure 2. The results indicated that the blood concentrations rapidly increased after administration, with peak values observed on Day 6 post-administration. The peak concentrations for the 150mg groups were 23.75 µg/mL The blood concentrations remained relatively stable within Day 29. By Day 183, the blood concentrations had decreased to 6.918 µg/mL (Figure 2). The half-life of SA55 was 103 days. Table1 summarized the main PK parameters of SA55 injection with 150mg single dose.

**Figure 2.**
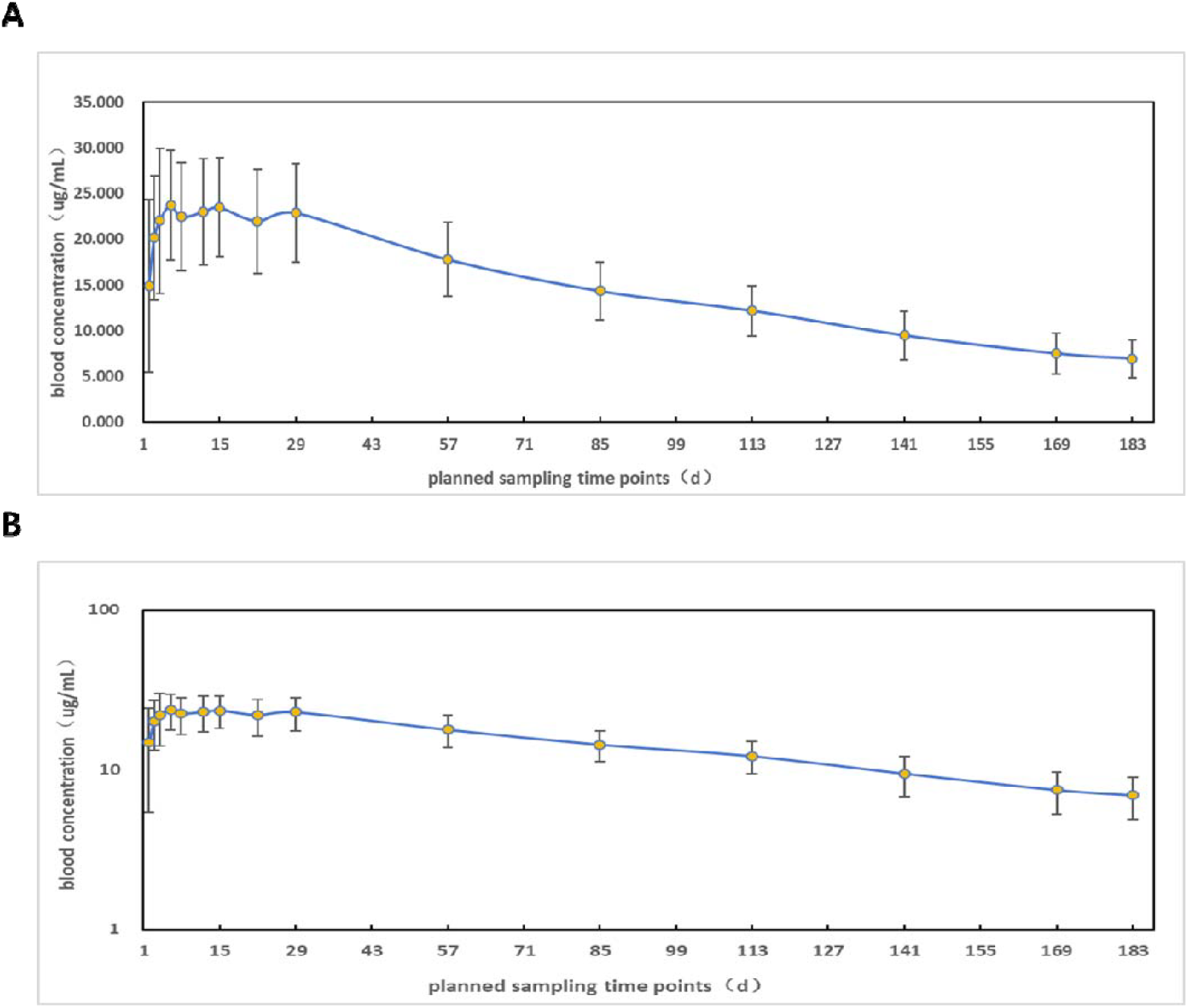
Pharmacokinetics characteristics (PKCS). Average drug concentration-time plot after administration of 150mg dose groups (A: linear scale, B: semi-logarithmic scale).

**Table 1.**
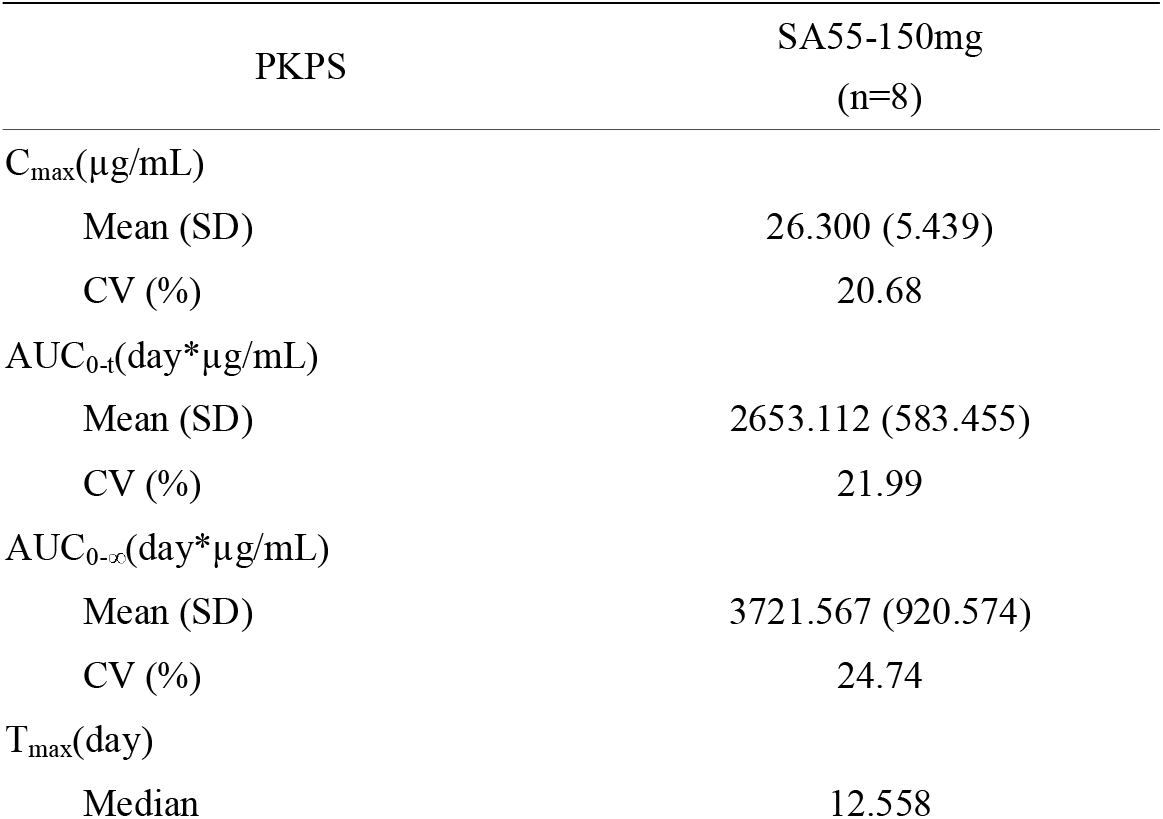
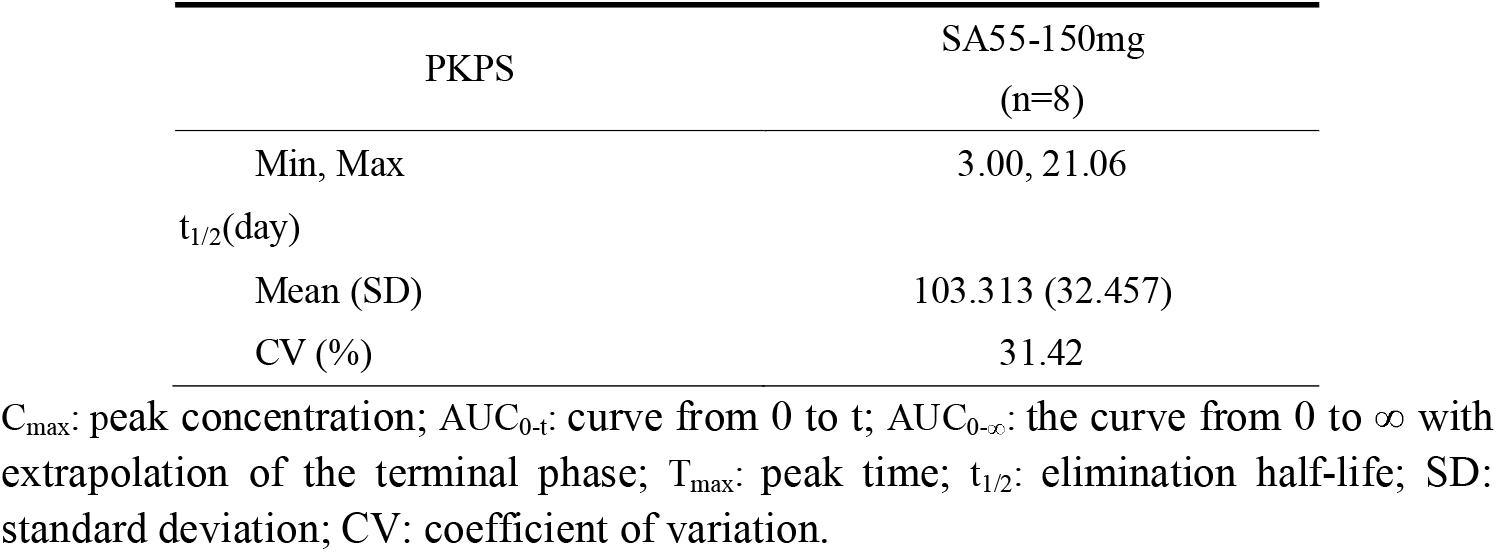
Summary of pharmacokinetic parameters (PKPS)

| PKPS | SA55-150mg<br>(n=8) |
| --- | --- |
| $C_{max}(\mu\text{g/mL})$ | |
| Mean (SD) | 26.300 (5.439) |
| CV (%) | 20.68 |
| $AUC_{0-t}(\text{day} \cdot \mu\text{g/mL})$ | |
| Mean (SD) | 2653.112 (583.455) |
| CV (%) | 21.99 |
| $AUC_{0-\infty}(\text{day} \cdot \mu\text{g/mL})$ | |
| Mean (SD) | 3721.567 (920.574) |
| CV (%) | 24.74 |
| $T_{max}(\text{day})$ | |
| Median | 12.558 |
| Min, Max | 3.00, 21.06 |
| $t_{1/2}$(day) | |
| Mean (SD) | 103.313 (32.457) |
| CV (%) | 31.42 |

$C_{max}$ : peak concentration; $AUC_{0-t}$ : curve from 0 to t; $AUC_{0-\infty}$ : the curve from 0 to ∞ with extrapolation of the terminal phase; $T_{max}$ : peak time; $t_{1/2}$ : elimination half-life; SD: standard deviation; CV: coefficient of variation.

## Discussion

This article reported the serum PK characteristics of the SA55 injection with 150mg single dose in healthy participants.

The serum PK characteristics of 150mg group indicated a T_max_ approximately 12.6 days. However, the serum concentration increased rapidly 1 day after the administration of SA55 injection, exceeded half of the Cmax, and reached a level approximating the C_max_ by 3 days after administration. This result indicates that the administration of SA55 injection is capable of rapidly achieving elevated plasma levels, which may confer prophylactic or therapeutic benefits. Furthermore, the half-life of SA55 injection exceeds three months, indicated its potential utility as a pre-exposure prophylactic agent against COVID-19, particularly for special populations who are ineligible for or have contraindications to COVID-19 vaccines.

## Conclusion

The PK results indicated that the blood concentration of SA55 increased rapidly, with a half-life exceeds 3 months, suggesting that SA55 injection can provide rapid and long-term protection against COVID-19. This implied that SA55 injection has the potential to serve as treatment and long-acting preventive antibody against COVID-19.

## Data Availability

All data produced in the present study are available upon reasonable request to the authors

## Author Contributions

X.M., C.H. and R.J. designed and generated the study protocol. Y.C. provided oversight and supervision. Y.Z., J.W., C.H. and R.J. coordinated the trial and responsible for the field work. J.L. conducted the neutralization assays. Z.Y. and J.W. wrote the original draft of the manuscript. X.M., G.Z., J.L. and Y.C. reviewed and revised the manuscript. All authors had access to the data in the study and had final responsibility for the decision to submit for publication.

## Acknowledgements

We thank all the participants for their contributions to this study, as well as the doctors and nurses who participated in this study.

## Informed Consent Statement

Informed consent was obtained from all participants involved in the study.

